# Real-world effectiveness of NVX-CoV2373 COVID-19 vaccine in immunocompromised individuals in South Korea

**DOI:** 10.1101/2025.04.24.25326359

**Authors:** Eunseon Gwak, Seung-Ah Choe, Kyuwon Kim, Erdenetuya Bolormaa, Manuela H. Gschwend, Jonathan Fix, Muruga Vadivale, Matthew D. Rousculp, Young June Choe

**Affiliations:** Department of Preventive Medicine, Korea University College of Medicine, Seoul, Korea; Research and Management Center for Health Risk of Particulate Matter, Korea University, Seoul, Korea; Novavax, Inc., Gaithersburg, MD, USA

**Author notes:** Equally contributed to the manuscript (Eunseon Gwak, Manuela H. Gschwend, Matthew D. Rousculp, Young June Choe). **Co-Correspondence**: Manuela H. Gschwend, PhD, Novavax, Inc., Gaithersburg, MD 20878, USA,; and Young June Choe, MD, PhD. Department of Pediatrics, Korea University College of Medicine, 73 Goryeodae-ro, Seongbuk-gu, Seoul 02841, Republic of Korea.

**Keywords:** COVID-19 vaccines, vaccination, heterologous, NVX-CoV2373, immunocompromised

## Abstract

**Background:** NVX-CoV2373 is a nanoparticle, protein-based COVID-19 vaccine. Individuals who are immunocompromised (IIC) are at high risk for infection and severe disease; however, real-world NVX-CoV2373 effectiveness data in IIC are limited.

**Methods:** South Korean IIC aged ≥12 years who received a primary series, third dose, or fourth dose of NVX-CoV2373 were identified in The Korea Disease Control and Prevention Agency-COVID-19-National Health Insurance Service (K-COV-N) database. IIC were propensity score matched to non-immunocompromised (non-IC) individuals to minimize potential confounding. Outcomes were any and severe SARS-CoV-2 infections, collected in cumulative 30-day risk windows through 180 days post vaccination in primary series and third and fourth dose groups. Adjusted hazard ratios (aHRs) measured relative vaccine effectiveness by comparing IIC and non-IC individuals across dose groups, overall, and by immunocompromising condition.

**Results:** A total of 755,727 doses of NVX-CoV2373 were administered to IIC February– December 2022, with 403,259 IIC included in this analysis. Through 180 days, aHRs (95% CI) for any SARS-CoV-2 infection were 1.10 (1.06–1.14), 1.05 (1.01–1.09), and 1.03 (1.02–1.05) for the primary series, third-dose, and fourth-dose groups; severe infection: 0.76 (0.52–1.12), 0.90 (0.53–1.51), and 1.11 (0.87–1.41), respectively. Risk estimates for any infection were relatively consistent across risk windows and among most immunocompromising conditions.

**Conclusion:** NVX-CoV2373 provided similar protection among IIC and non-IC individuals regardless of dose administered and IC condition.

## Introduction

Emerging in 2019, SARS-CoV-2 triggered the global COVID-19 pandemic. Despite announcement of the end of the pandemic in May 2023, the ongoing evolution of new strains continues to pose a substantial global health threat [1]. The evolution of immune-evasive variants has underscored the necessity for ongoing vaccine development and adaptation [2,3]. While various vaccines have been authorized/approved for use, their effectiveness in specific populations, particularly individuals who are immunocompromised (IIC), remains a critical area of investigation based on the increased susceptibility to infection and progression to severe/critical disease among IIC [4].

IIC comprise 2% to 3% of the overall population and are a heterogenous group of individuals with genetic or acquired conditions that disrupt normal immune function and/or are taking essential immune-modifying therapies after certain procedures (e.g., organ transplants) or to control disease [5,6]. Higher rates of COVID-19–related hospitalizations in IIC [4,7] defined the group as an at-risk population, supporting their eligibility in South Korea to receive biannual vaccinations (versus a single seasonal dose) for additional protection against infection/disease [8].

IIC are often excluded from or vastly underrepresented in clinical trials of vaccines, and meta-analyses and systematic reviews of COVID-19 vaccine clinical trials have mostly focused on mRNA-based COVID-19 vaccines [9–12]. Because of altered immune responses, IIC may not elicit as robust responses to a vaccine as non-immunocompromised (non-IC) individuals, having presented with lower seroconversion rates [13] and vaccine effectiveness (VE) [14]. For that reason, understanding the real-world effectiveness of different vaccines in IIC is paramount for optimizing vaccination strategies and protecting this high-risk group.

The Novavax COVID-19 vaccine is a nanoparticle vaccine expressing recombinant SARS-CoV-2 spike (rS) protein with Matrix-M™ adjuvant. A formulation targeting ancestral SARS-CoV-2 (NVX-CoV2373) was authorized for use in South Korea in 2022 and has since been amended for use of updated formulations targeting relevant variant strains (NVX-CoV2601 [XBB.1.5]; NVX-CoV2705 [JN.1]) [15].

This study assesses the comparative effectiveness of NVX-CoV2373 in preventing SARS-CoV-2 infection and severe illness among IIC versus non-IC individuals in the general population. By utilizing population-based data from the period when Omicron variants (e.g., BA.1/1.1, BA.2/2.3, and BA.5/5.2) [16] predominated, we seek to provide valuable real-world insights into the vaccine’s performance in relation to immunocompromised status in South Korea. The results of this study will inform evidence-based decision-making regarding vaccination policies and public health strategies.

## Materials and methods

### Data source and study population

The Korea Disease Control and Prevention Agency-COVID-19-National Health Insurance Service (K-COV-N) cohort database was utilized for this study. This database integrates COVID-19 vaccination records, SARS-CoV-2 infection surveillance data, and health insurance claims data, providing a comprehensive view of the patient population. This study focused on individuals aged ≥12 years who had received at least one dose of the NVX-CoV2373 vaccine in South Korea between February 1 and December 31, 2022. These individuals were required to have at least 365 days of observation in the database prior to their vaccination date. Individuals with mixed primary series vaccinations (i.e., having received first and second doses of COVID-19 vaccines from different manufacturers) were excluded.

The IIC population was identified based on the presence of specific International Classification of Diseases, 10^th^ revision (ICD-10) codes, including conditions such as rheumatologic or inflammatory disorder (L40–L45, M05–M14, G35, K50–K51); metabolic disorder (E10–E13; K90–K90.4); solid organ malignancy (C18, C22, C25, C34, C50, C53, C56, C64); chronic kidney disease (N18.1–6, N18.9); hematologic malignancy (C81–C96); organ or stem cell transplant (Z94.0–Z94.9); other intrinsic immune condition or immunodeficiency (D80–D84, D71, D73, Z90.81); HIV/AIDS (B20–B24) (**Table 1**).

**Table 1:** Types of immunocompromising conditions included in this study.

| Condition category | Condition name (ICD-10 code) | Study participants, n (%) |
| --- | --- | --- |
| Rheumatologic/ inflammatory disorder | Psoriasis (L40) | 178,205 (56.0) |
|  | Parapsoriasis (L41) |  |
|  | Pityriasis rosea (L42) |  |
|  | Lichen planus (L43) |  |
|  | Other papulosquamous disorders (L44) |  |
|  | Other specified erythematous conditions (L45) |  |
|  | Rheumatoid arthritis with rheumatoid factor (M05) |  |
|  | Other rheumatoid arthritis (M06) |  |
|  | Psoriatic and enteropathic arthropathies (M07) |  |
|  | Juvenile arthritis (M08)<br>Juvenile arthritis in diseases classified elsewhere (M09)<br>Gout (M10)<br>Other crystal arthropathies (M11)<br>Other specific arthropathies (M12)<br>Other arthritis, not elsewhere classified (M13)<br>Secondary arthritis (M14)<br>Multiple sclerosis (G35)<br>Crohn's disease (K50)<br>Ulcerative colitis (K51) |  |
| Metabolic disorder | Type 1 diabetes mellitus (E10)<br>Type 2 diabetes mellitus (E11)<br>Malnutrition-related diabetes mellitus (E12)<br>Other specified diabetes mellitus (E13)<br>Intestinal malabsorption (K90)<br>Celiac disease (K90.0)<br>Tropical sprue (K90.1)<br>Blind loop syndrome (K90.2)<br>Pancreatic steatorrhea (K90.3)<br>Other intestinal malabsorption (K90.4) | 177,691 (55.9) |
| Solid organ malignancy | Malignant neoplasm of the colon (C18)<br>Malignant neoplasm of the liver and intrahepatic bile ducts (C22)<br>Malignant neoplasm of the pancreas (C25)<br>Malignant neoplasm of the bronchus and lung (C34)<br>Malignant neoplasm of the breast (C50)<br>Malignant neoplasm of the cervix uteri (C53)<br>Malignant neoplasm of the ovary (C56)<br>Malignant neoplasm of the kidney, except the renal pelvis (C64) | 24,818 (7.8) |
| Chronic kidney disease | Chronic kidney disease, Stage 1 (N18.1)<br>Chronic kidney disease, Stage 2 (N18.2)<br>Chronic kidney disease, Stage 3 (N18.3)<br>Chronic kidney disease, Stage 4 (N18.4)<br>Chronic kidney disease, Stage 5 (N18.5)<br>End-stage renal disease (N18.6)<br>Chronic kidney disease, unspecified (N18.9) | 14,700 (4.6) |
| Hematologic malignancy | Hodgkin lymphoma (C81)<br>Follicular lymphoma (C82)<br>Non-follicular lymphoma (C83)<br>Mature T/NK-cell lymphomas (C84)<br>Other and unspecified types of non-Hodgkin lymphoma (C85)<br>Other specified types of T/NK-cell lymphoma (C86)<br>Malignant immunoproliferative diseases and certain other B-cell lymphomas (C88)<br>Multiple myeloma and malignant plasma cell neoplasms (C90)<br>Lymphoid leukemia (C91)<br>Myeloid leukemia (C92)<br>Monocytic leukemia (C93)<br>Other leukemias of specified cell type (C94)<br>Leukemia of unspecified cell type (C95) | 2,354 (0.7) |
|  | Other and unspecified malignant neoplasms of lymphoid, hematopoietic, and related tissue (C96) |  |
| Organ or stem cell transplant | Kidney transplant status (Z94.0)<br>Heart transplant status (Z94.1)<br>Lung transplant status (Z94.2)<br>Heart and lungs transplant status (Z94.3)<br>Liver transplant status (Z94.4)<br>Skin transplant status (Z94.5)<br>Bone transplant status (Z94.6)<br>Corneal transplant status (Z94.7)<br>Other transplanted organ and tissue status (Z94.8)<br>Transplanted organ and tissue status, unspecified (Z94.9) | 1682 (0.5) |
| Other intrinsic immune condition/immunodeficiency | Immunodeficiency with predominantly antibody defects (includes conditions such as X-linked agammaglobulinemia and common variable immunodeficiency; D80)<br>Combined immunodeficiencies (e.g., severe combined immunodeficiency; D81)<br>Immunodeficiency associated with other major defects (e.g., DiGeorge syndrome; D82)<br>Common variable immunodeficiency (D83)<br>Other immunodeficiencies (e.g., selective IgA deficiency; D84)<br>Functional disorders of polymorphonuclear neutrophils (D71)<br>Hyposplenism (D73.0)<br>Chronic congestive splenomegaly (D73.1)<br>Abscess of the spleen (D73.2)<br>Splenic disease, unspecified (D73.9)<br>Acquired absence of spleen (Z90.81) | 593 (0.2) |
| HIV/AIDS | HIV disease resulting in infectious and parasitic diseases (e.g., opportunistic infections such as tuberculosis, candidiasis, or pneumocystis pneumonia due to HIV (B20)<br>HIV disease resulting in malignant neoplasms (e.g., cancers commonly associated with HIV, like Kaposi's sarcoma or non-Hodgkin's lymphoma (B21)<br>HIV disease resulting in other specified diseases (e.g., encephalopathy or wasting syndrome linked to HIV (B22)<br>HIV disease resulting in other conditions (B23)<br>Unspecified HIV disease (e.g., used when HIV is present but without further specification of associated conditions; B24) | 392 (0.1) |

### Outcomes

The primary outcomes of interest were any SARS-CoV-2 infection and severe SARS-CoV-2 infection. Severe infection was defined as either admission to an intensive care unit (ICU) due to SARS-CoV-2 infection or death following SARS-CoV-2 infection. ICU admissions were identified using specific treatment codes in conjunction with a primary or secondary diagnosis of conditions related to SARS-CoV-2 infection (ICD-10 codes: B342, B972, Z208, Z209, Z115, U181, Z038, U071, U072, U08, U09, and U10), as previously described [17]. Deaths occurring within 8 weeks of a confirmed SARS-CoV-2 infection were considered COVID-19–related, given the lack of precise cause-of-death data. A sensitivity analysis was performed with data from a subset of participants considered severely immunocompromised, defined as those with records indicating solid organ malignancy; hematologic malignancy; rheumatologic or inflammatory disorder; or other intrinsic immune condition or immunodeficiency.

### Statistical analysis

Our analyses were stratified by the dose of NVX-CoV2373 (i.e., primary series, third dose, or fourth dose), and descriptive baseline characteristics and demographics were provided for each group. Propensity score matching (1:1 match) was employed to balance baseline characteristics between enrolled IIC and a matched general population without an immunocompromised condition (i.e., non-IC) to minimize potential confounding. Balance was evaluated using absolute standardized difference (aSD) where an aSD >0.1 indicated a significant difference. Exact matching was completed on the year-month of vaccination. Relative vaccine effectiveness (rVE) was estimated using Cox proportional hazard models to produce adjusted hazard ratios (aHRs) and corresponding 95% CIs considering time-varying vaccination status. Date of NVX-CoV2373 vaccination was considered the index date (date of a second dose for the primary series). For the primary series group, the follow-up period for each individual began 14 days after the index date; for the analyses of third and fourth doses, the follow-up began 7 days after the index date. Follow-up continued through another 180 days, or until the earliest of the following events: documentation of SARS-CoV-2 infection, death, administration of a subsequent COVID-19 vaccine dose, or the end of the study period (December 31, 2022). The risk windows for evaluating outcomes were in 30-day intervals at 30, 60, 90, 120, 150, and 180 days after the start of follow-up. Regression models included adjustment for age at vaccination, sex, living in the non-capital area, presence of disability, employment, level of income, Charlson comorbidity index (CCI ≥3), obesity (body-mass index >30 kg/m^2^), current smoker, and SARS-CoV-2 infection within 6 months prior to the index date.

A negative control analysis calculating adjusted odds ratios (aORs) and 95% CIs for any SARS-CoV-2 infection and severe infection during 0–5 days post vaccination was performed using logistic regression to assess residual confounding in VE estimates. All statistical analyses were performed using SAS 9.4^®^ software (SAS Institute, Cary, NC, USA), and the study protocol was approved by the Korea University Institutional Review Board (IRB No. 2023AN0124).

## Results

### Study populations

A total of 755,727 doses of NVX-CoV2373 were administered to IIC as homologous primary or homologous or heterologous third and fourth doses between February 1, 2022, and December 31, 2022 (**Figure 1**). A total of 403,259 IIC met inclusion criteria, with the main reasons for exclusion being non-homologous primary series and vaccination other than NVX-CoV2373. The immunocompromising conditions with the highest proportions of enrolled IIC were rheumatologic/inflammatory (56.0%) and metabolic (55.9%) disorders (**Table 1**).

**Figure 1.**
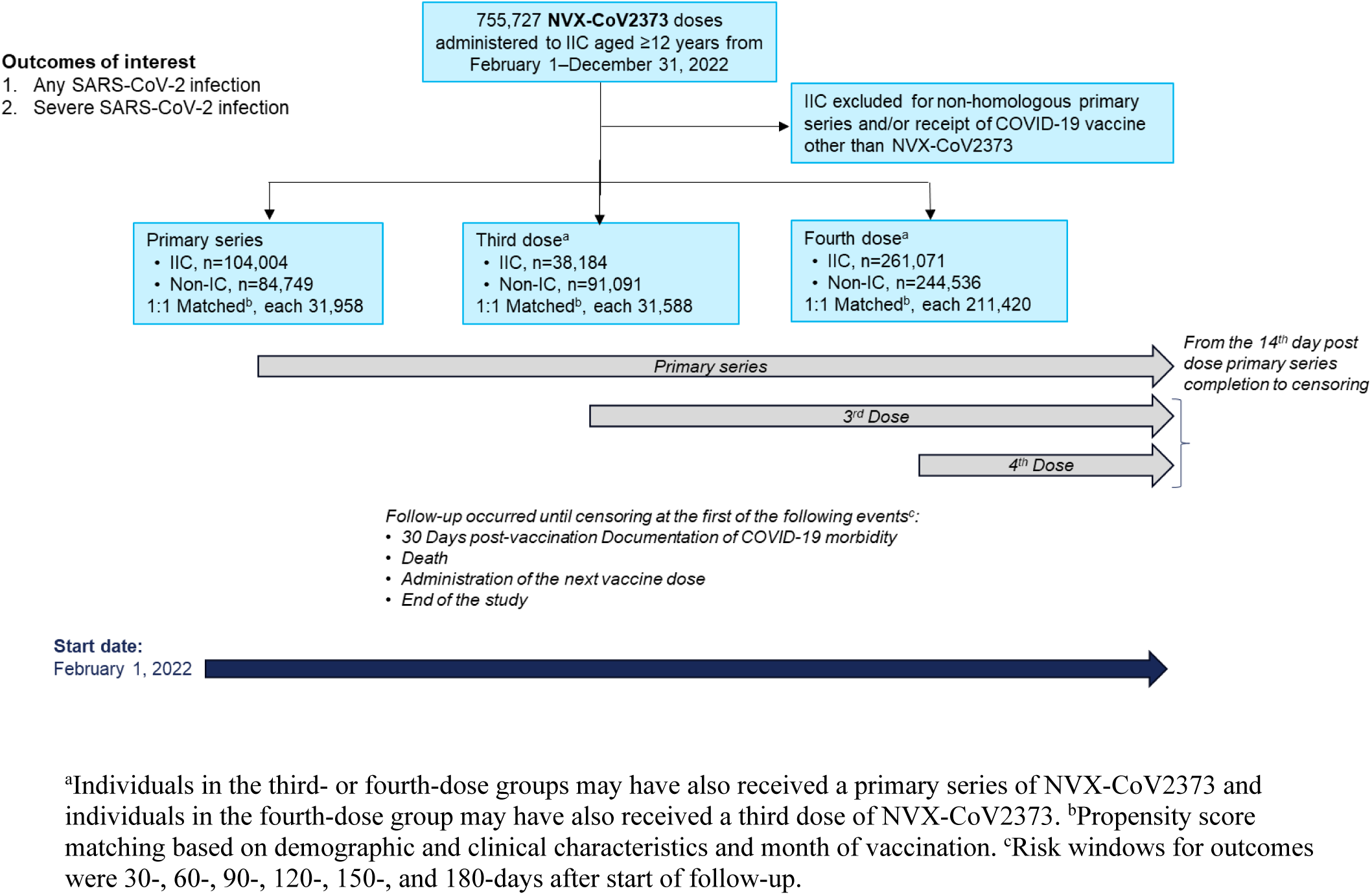
Study design for relative vaccine effectiveness comparing IIC and non-IC recipients of NVX-CoV2373. IC, immunocompromised; IIC, individuals who are immunocompromised.

Descriptive statistics showed that IIC, compared to the non-IC group, tended to be older, were more likely to be living in the non-capital area, had comorbidities or disability, were unemployed, and were medical aid beneficiaries (low/no income), whether they received NVX-CoV2373 as the primary series, third dose, or fourth dose (**Table 2**). After propensity score matching, the IIC and non-IC groups each had 31,958 participants in the primary series groups, 31,588 in the third-dose groups, and 211,420 participants in the fourth-dose groups (**Figure 1**; **Table 2**). There was a significantly greater proportion of matched IIC compared with non-IC individuals who were aged 70–79 years and a significantly smaller proportion who were employed among recipients of a fourth dose (**Table 2**). No significant differences were observed between groups among primary series and third-dose recipients.

**Table 2.** Participant baseline characteristics by dose group in IIC and non-IC individuals, before and after propensity score matching.

| DOSE GROUP<br>Variable, n/N (%) | Before matching |  |  | After matching |  |  |
| --- | --- | --- | --- | --- | --- | --- |
|  | IIC | Non-IC | *P* | IIC | Non-IC | aSD<sup>a</sup> |
| **PRIMARY SERIES** |  |  |  |  |  |  |
| N | 38,096 | 84,749 |  | 31,958 | 31,958 |  |
| Age |  |  |  |  |  |  |
| 12–17 | 42 (0.1) | 575 (0.7) | <0.001 | 42 (0.1) | 42 (0.1) | 0.000 |
| 18–24 | 1687 (4.4) | 12,374 (14.6) |  | 1686 (5.3) | 1639 (5.1) | 0.005 |
| 25–39 | 5076 (13.3) | 25,044 (29.6) |  | 5042 (15.8) | 4879 (15.3) | 0.013 |
| 40–49 | 6669 (17.5) | 18,558 (21.9) |  | 6198 (19.4) | 6175 (19.3) | 0.002 |
| 50–59 | 6482 (17.0) | 10,834 (12.8) |  | 5519 (17.3) | 5467 (17.1) | 0.005 |
| 60–69 | 7921 (20.8) | 8453 (10.0) |  | 6368 (19.9) | 6365 (19.9) | 0.000 |
| 70–79 | 5196 (13.6) | 3814 (4.5) |  | 3746 (11.7) | 3627 (11.4) | 0.013 |
| ≥80 | 5023 (13.2) | 5097 (6.0) |  | 3357 (10.5) | 3764 (11.8) | 0.044 |
| Sex |  |  |  |  |  |  |
| Male | 16,895 (44.4) | 36,831 (43.5) | 0.004 | 14,109 (44.2) | 14,107 (44.1) | 0.000 |
| Female | 21,201 (55.7) | 47,918 (56.5) |  | 17,849 (55.9) | 17,851 (55.9) |  |
| Non-capital area<sup>b</sup> | 20,257 (53.2) | 42,187 (49.8) | <0.001 | 16,656 (52.1) | 16,614 (52.0) | 0.003 |
| CCI ≥3 | 7745 (20.3) | 1432 (1.7) | <0.001 | 1620 (5.1) | 1432 (4.5) | 0.020 |
| Disability | 5349 (14.0) | 5178 (6.1) | <0.001 | 3981 (12.5) | 3682 (11.5) | 0.031 |
| Employed | 9673 (25.4) | 29,305 (34.6) | <0.001 | 8953 (28.0) | 8839 (27.7) | 0.008 |
| Income level<sup>c</sup> |  |  |  |  |  |  |
| Medical aid beneficiary | 3186 (8.6) | 3184 (3.9) | <0.001 | 2697 (8.7) | 2395 (7.7) | 0.040 |
| 1Q | 5090 (13.7) | 12,054 (14.7) |  | 4204 (13.5) | 4229 (13.5) | 0.002 |
| 2Q | 6629 (17.8) | 16,653 (20.3) |  | 5697 (18.3) | 5754 (18.4) | 0.005 |
| 3Q | 8425 (22.7) | 21,209 (25.9) |  | 7033 (22.6) | 7095 (22.7) | 0.005 |
| 4Q | 13,873 (37.3) | 28,797 (35.2) |  | 11,540 (37.0) | 11,167 (37.7) | 0.015 |
| Obesity (BMI >30kg/m<sup>2</sup>) | 965 (2.5) | 1429 (1.7) | <0.001 | 897 (2.8) | 729 (2.3) | 0.037 |
| Current smoking | 1799 (4.7) | 3769 (4.5) | 0.032 | 1667 (5.2) | 1585 (5.0) | 0.012 |
| Prior SARS-CoV-2 infection within 6 months | 4993 (13.1) | 10,670 (12.6) | 0.012 | 4119 (12.9) | 3977 (12.4) | 0.013 |
| **THIRD DOSE** |  |  |  |  |  |  |
| N | 36,184 | 91,091 |  | 31,588 | 31,588 |  |
| Age |  |  |  |  |  |  |
| 12–17 | 46 (0.1) | 516 (0.6) | <0.001 | 46 (0.2) | 45 (0.1) | 0.001 |
| 18–24 | 2359 (6.5) | 18,724 (20.6) |  | 2356 (7.5) | 2339 (7.4) | 0.002 |
| 25–39 | 4386 (12.1) | 22,192 (24.4) |  | 4349 (13.8) | 4259 (13.5) | 0.007 |
| 40–49 | 7265 (20.1) | 21,668 (23.8) |  | 6873 (21.8) | 6861 (21.7) | 0.001 |
| 50–59 | 8208 (22.7) | 14,748 (16.2) |  | 7291 (23.1) | 7297 (23.1) | 0.000 |
| 60–69 | 7789 (21.5) | 8531 (9.4) |  | 6338 (20.1) | 6547 (20.7) | 0.019 |
| 70–79 | 3834 (10.6) | 2565 (2.8) |  | 2758 (8.7) | 2520 (8.0) | 0.030 |
| ≥80 | 2297 (6.4) | 2147 (2.4) |  | 1577 (5.0) | 1720 (5.5) | 0.022 |
| Sex |  |  |  |  |  |  |
| Male | 16,318 (45.1) | 41,155 (45.2) | 0.789 | 14,311 (45.3) | 14,430 (45.7) | 0.08 |
| Female | 19,866 (54.9) | 49,936 (54.8) |  | 17,277 (54.7) | 17,158 (54.3) |  |
| Non-capital area<sup>b</sup> | 17,139 (47.4) | 39,719 (43.6) | <0.001 | 14,572 (46.1) | 14,606 (46.2) | 0.002 |
| CCI ≥3 | 5611 (15.5) | 941 (1.0) | <0.001 | 1031 (3.3) | 940 (3.0) | 0.011 |
| Disability | 3306 (9.1) | 3155 (3.5) | <0.001 | 2592 (8.2) | 2288 (7.2) | 0.040 |
| Employed | 13,712 (37.9) | 43,356 (47.6) | <0.001 | 12,798 (40.5) | 12,931 (40.9) | 0.009 |
| Income level<sup>c</sup> |  |  |  |  |  |  |
| Medical aid beneficiary | 2120 (6.0) | 1978 (2.2) | <0.001 | 1615 (5.2) | 1430 (4.6) | 0.030 |
| 1Q | 4584 (12.9) | 11,480 (12.8) |  | 3988 (12.9) | 3992 (12.9) | 0.000 |
| 2Q | 6303 (17.7) | 17,804 (19.9) |  | 5634 (18.2) | 5709 (18.4) | 0.006 |
| 3Q | 7881 (22.2) | 22,179 (24.8) |  | 6993 (22.6) | 6974 (22.5) | 0.001 |
| 4Q | 14,671 (41.3) | 36,059 (40.3) |  | 12,783 (41.2) | 12,918 (41.6) | 0.009 |
| Obesity (BMI > 30kg/m<sup>2</sup>) | 1165 (3.2) | 1862 (2.0) | <0.001 | 1072 (3.4) | 1038 (3.3) | 0.007 |
| Current smoking | 2180 (6.0) | 5468 (6.0) | 0.882 | 2050 (6.5) | 1952 (6.2) | 0.013 |
| Prior SARS-CoV-2 infection within 6 months | 4964 (13.7) | 12,652 (13.9) | 0.427 | 4266 (13.5) | 4087 (12.9) | 0.016 |
| **FOURTH DOSE** |  |  |  |  |  |  |
| N | 261,071 | 244,536 |  | 211,420 | 211,420 |  |
| Age |  |  |  |  |  |  |
| 12–17 | 0 (0.0) | 0 (0.0) | <0.001 | 0 (0.0) | 0 (0.0) |  |
| 18–24 | 292 (0.1) | 1016 (0.4) |  | 289 (0.1) | 390 (0.2) | 0.009 |
| 25–39 | 883 (0.3) | 2043 (0.8) |  | 832 (0.4) | 1082 (0.5) | 0.015 |
| 40–49 | 3025 (1.2) | 3898 (1.6) |  | 2725 (1.3) | 2820 (1.3) | 0.004 |
| 50–59 | 33,560 (12.9) | 53,034 (21.7) |  | 30,751 (14.5) | 35,650 (16.9) | 0.062 |
| 60–69 | 111,135 (42.6) | 106,268 (43.5) |  | 94,513 (44.7) | 104,341 (49.4) | 0.094 |
| 70–79 | 79,078 (30.3) | 54,289 (22.2) |  | 58,426 (27.6) | 43,931 (20.8) | 0.156 |
| ≥80 | 33,098 (12.7) | 23,988 (9.8) |  | 23,884 (11.3) | 23,206 (11.0) | 0.010 |
| Sex |  |  |  |  |  |  |
| Male | 127,945 (49.0) | 123,957 (50.7) | <0.001 | 104,367 (49.4) | 111,121 (52.6) | 0.064 |
| Female | 133,126 (51.0) | 120,579 (49.3) |  | 107,053 (50.6) | 100,299 (47.4) |  |
| Non-capital area<sup>b</sup> | 109,693 (58.0) | 104,534 (57.3) | <0.001 | 121,640 (57.5) | 122,218 (57.8) | 0.006 |
| CCI ≥3 | 52,537 (20.1) | 7993 (3.3) | <0.001 | 8008 (3.8) | 7991 (3.8) | 0.000 |
| Disability | 36,704 (14.1) | 24,420 (10.0) | <0.001 | 24,927 (11.8) | 21,042 (10.0) | 0.057 |
| Employed | 64,931 (24.9) | 78,871 (32.3) | <0.001 | 57,770 (27.3) | 73,458 (34.8) | 0.165 |
| Income level<sup>c</sup> |  |  |  |  |  |  |
| Medical aid beneficiary | 17,513 (6.8) | 10,844 (4.5) | <0.001 | 11,307 (5.4) | 8561 (4.1) | 0.057 |
| 1Q | 38,438 (14.9) | 37,314 (15.5) |  | 31,648 (15.2) | 32,521 (15.6) | 0.012 |
| 2Q | 48,735 (18.9) | 48,652 (20.2) |  | 40,749 (19.5) | 42,298 (20.3) | 0.019 |
| 3Q | 53,552 (20.8) | 51,903 (21.5) |  | 44,330 (21.3) | 46,224 (22.2) | 0.022 |
| 4Q | 99,364 (38.6) | 92,520 (38.4) |  | 80,568 (38.6) | 78,655 (37.8) | 0.019 |
| Obesity (BMI> 30kg/m<sup>2</sup>) | 7528 (2.9) | 4963 (2.0) | <0.001 | 5576 (2.6) | 3703 (1.8) | 0.057 |
| Current smoking | 13,916 (5.3) | 14,772 (6.0) | <0.001 | 11,643 (5.5) | 13,020 (6.2) | 0.028 |
| Prior SARS-CoV-2 infection within 6 months | 55,186 (21.1) | 50,258 (20.6) | <0.001 | 43,552 (20.6) | 40,691 (19.3) | 0.033 |

<sup>a</sup>aSD values >0.1 indicate a significant imbalance between IIC and non-IC groups. <sup>b</sup>Indication of whether participants were living inside or outside of the Seoul Capital Area. <sup>c</sup>Income was categorized as medical aid beneficiary (no/lowest income) and then into quartiles of 10–40%, >40–60%, >60–80%, and >80–100%. BMI, body-mass index; CCI, Charlson Comorbidity Index; IC, immunocompromised; IIC, individuals who are immunocompromised; aSD, absolute standardized difference.

### Incidence of SARS-CoV-2 infection

Infection incidence rate (IR) per 1000 person-days was determined among the IIC and non-IC populations across the defined risk windows. Among primary series recipients, the IR of any SARS-CoV-2 infection within the 30-day risk window (44–74 days post-vaccination) was 1.39 (95% CI: 1.32–1.47) for the IIC group and 1.32 (95% CI: 1.25–1.40) for the non-IC group (**Table 3**). During the same period, the IR (95% CI) for severe infection was 0.02 (0.01–0.03) for the IIC group and 0.02 (0.02–0.04) for the non-IC group. Through the 180-day risk window, the IR (95% CI) for any SARS-CoV-2 infection was 1.21 (1.18–1.24) and 1.10 (1.07–1.13) for the IIC and non-IC groups, respectively, and 0.01 (0.01–0.01) in both groups for severe SARS-CoV-2 infection.

**Table 3.** The IRs and aHRs of any and severe SARS-CoV-2 infection after receipt of homologous primary series or third or fourth COVID-19 vaccine dose of NVX-CoV2373 in IIC and non-IC individuals.

| Risk window | Dose SARS-CoV-2 infection | IIC<sup>a</sup> |  |  | Non-IC |  |  | aHR (95% CI)<sup>c</sup> |
| --- | --- | --- | --- | --- | --- | --- | --- | --- |
|  |  | Events, n | Time<sup>b</sup> | IR (95% CI)<sup>b</sup> | Events, n | Time<sup>b</sup> | IR (95% CI)<sup>b</sup> |  |
| 30 days | **Primary series** |  |  |  |  |  |  |  |
|  | Any | 1371 | 933 | 1.39 (1.32–1.47) | 1306 | 934 | 1.32 (1.25–1.40) | 1.04 (0.96–1.12) |
|  | Severe | 16 | 958 | 0.02 (0.01–0.03) | 23 | 958 | 0.02 (0.02–0.04) | 0.72 (0.37–1.38) |
|  | **Third** |  |  |  |  |  |  |  |
|  | Any | 2088 | 913 | 2.19 (2.10–2.29) | 2073 | 913 | 2.19 (2.10–2.29) | 1.01 (0.95–1.08) |
|  | Severe | 11 | 947 | 0.01 (0.01–0.02) | 6 | 948 | 0.01 (0.00–0.01) | 1.69 (0.61–4.66) |
|  | **Fourth** |  |  |  |  |  |  |  |
|  | Any | 5086 | 6256 | 0.78 (0.76–0.80) | 4989 | 6259 | 0.76 (0.74–0.79) | 1.10 (1.06–1.15) |
|  | Severe | 27 | 6342 | 0.00<br>(0.00–0.01) | 22 | 6342 | 0.00<br>(0.00–0.01) | 1.14<br>(0.64–2.04) |
| **60 days** | **Primary series** |  |  |  |  |  |  |  |
|  | Any | 1811 | 1841 | 0.94<br>(0.90–0.99) | 1701 | 1845 | 0.88<br>(0.84–0.93) | 1.06<br>(0.99–1.13) |
|  | Severe | 19 | 1917 | 0.01<br>(0.01–0.02) | 36 | 1916 | 0.02<br>(0.01–0.03) | 0.55<br>(0.31–0.97) |
|  | **Third** |  |  |  |  |  |  |  |
|  | Any | 2975 | 1782 | 1.62<br>(1.56–1.68) | 2893 | 1784 | 1.58<br>(1.52–1.64) | 1.04<br>(0.99–1.09) |
|  | Severe | 18 | 1895 | 0.01<br>(0.01–0.02) | 15 | 1895 | 0.01<br>(0.00–0.01) | 1.08<br>(0.54–2.18) |
|  | **Fourth** |  |  |  |  |  |  |  |
|  | Any | 8076 | 12,398 | 0.63<br>(0.62–0.65) | 7884 | 12,405 | 0.62<br>(0.61–0.63) | 1.10<br>(1.06–1.13) |
|  | Severe | 59 | 12,684 | 0.00<br>(0.00–0.01) | 50 | 12,684 | 0.00<br>(0.00–0.01) | 1.09<br>(0.74–1.60) |
| **90 days** | **Primary series** |  |  |  |  |  |  |  |
|  | Any | 2275 | 2691 | 0.82<br>(0.78–0.85) | 2153 | 2703 | 0.77<br>(0.74–0.80) | 1.06<br>(1.00–1.12) |
|  | Severe | 23 | 2875 | 0.01<br>(0.01–0.01) | 41 | 2874 | 0.01<br>(0.01–0.02) | 0.58<br>(0.35–0.98) |
|  | **Third** |  |  |  |  |  |  |  |
|  | Any | 3439 | 2632 | 1.27<br>(1.23–1.32) | 3344 | 2636 | 1.24<br>(1.20–1.28) | 1.04<br>(0.99–1.09) |
|  | Severe | 19 | 2842 | 0.01<br>(0.00–0.01) | 18 | 2842 | 0.01<br>(0.00–0.01) | 0.95<br>(0.49–1.83) |
|  | **Fourth** |  |  |  |  |  |  |  |
|  | Any | 12,952 | 18,429 | 0.69<br>(0.68–0.70) | 12,643 | 18,445 | 0.67<br>(0.66–0.69) | 1.07<br>(1.04–1.10) |
|  | Severe | 82 | 19,024 | 0.00<br>(0.00–0.01) | 76 | 19,025 | 0.00<br>(0.00–0.01) | 1.02<br>(0.74–1.39) |
| **120 days** | **Primary series** |  |  |  |  |  |  |  |
|  | Any | 3419 | 3361 | 1.00<br>(0.96–1.03) | 3188 | 3393 | 0.92<br>(0.89–0.95) | 1.08<br>(1.03–1.13) |
|  | Severe | 32 | 3833 | 0.01<br>(0.01–0.01) | 52 | 3831 | 0.01<br>(0.01–0.02) | 0.64<br>(0.41–1.00) |
|  | **Third** |  |  |  |  |  |  |  |
|  | Any | 3962 | 3455 | 1.12<br>(1.09–1.16) | 3842 | 3465 | 1.09<br>(1.05–1.12) | 1.04<br>(1.00–1.09) |
|  | Severe | 24 | 3789 | 0.01<br>(0.00–0.01) | 23 | 3789 | 0.01<br>(0.00–0.01) | 0.94<br>(0.52–1.68) |
|  | **Fourth** |  |  |  |  |  |  |  |
|  | Any | 19,781 | 24,270 | 0.81<br>(0.80–0.82) | 19,300 | 24,300 | 0.79<br>(0.77–0.80) | 1.04<br>(1.02–1.06) |
|  | Severe | 111 | 25,364 | 0.00<br>(0.00–0.01) | 96 | 25,365 | 0.00<br>(0.00–0.00) | 1.08<br>(0.82–1.43) |
| **150 days** | **Primary series** |  |  |  |  |  |  |  |
|  | Any | 4739 | 3931 | 1.19<br>(1.15–1.22) | 4431 | 3990 | 1.09<br>(1.06–1.13) | 1.09<br>(1.04–1.13) |
|  | Severe | 42 | 4790 | 0.01<br>(0.01–0.01) | 60 | 4788 | 0.01<br>(0.01–0.02) | 0.74<br>(0.49–1.10) |
|  | **Third** |  |  |  |  |  |  |  |
|  | Any | 4747 | 4232 | 1.10<br>(1.07–1.13) | 4593 | 4251 | 1.06<br>(1.03–1.10) | 1.05<br>(1.00–10.9) |
|  | Severe | 29 | 4736 | 0.01<br>(0.00–0.01) | 27 | 4736 | 0.01<br>(0.00–0.01) | 0.97<br>(0.57–1.65) |
|  | **Fourth** |  |  |  |  |  |  |  |
|  | Any | 23,094 | 29,947 | 0.76<br>(0.75–0.77) | 22,441 | 29,997 | 0.74<br>(0.73–0.75) | 1.04<br>(1.02–1.06) |
|  | Severe | 131 | 31,703 | 0.00<br>(0.00–0.00) | 114 | 31,704 | 0.00<br>(0.00–0.00) | 1.08<br>(0.84–1.40) |
| **180 days** | **Primary series** |  |  |  |  |  |  |  |
|  | Any | 5459 | 4448 | 1.21<br>(1.18–1.24) | 5065 | 4542 | 1.10<br>(1.07–1.13) | 1.10<br>(1.06–1.14) |
|  | Severe | 47 | 5748 | 0.01<br>(0.01–0.01) | 64 | 5745 | 0.01<br>(0.01–0.01) | 0.76<br>(0.52–1.12) |
|  | **Third** |  |  |  |  |  |  |  |
|  | Any | 5452 | 4969 | 1.08<br>(1.05–1.11) | 5273 | 5000 | 1.04<br>(1.01–1.07) | 1.05<br>(1.01–1.09) |
|  | Severe | 29 | 5682 | 0.01<br>(0.00–0.01) | 29 | 5683 | 0.01<br>(0.00–0.01) | 0.90<br>(0.53–1.51) |
|  | **Fourth** |  |  |  |  |  |  |  |
|  | Any | 24,342 | 35,557 | 0.68<br>(0.67–0.69) | 23,626 | 35,631 | 0.66<br>(0.65–0.67) | 1.03<br>(1.02–1.05) |
|  | Severe | 150 | 38,041 | 0.00<br>(0.00–0.00) | 127 | 38,043 | 0.00<br>(0.00–0.00) | 1.11<br>(0.87–1.41) |

<sup>a</sup>Data are stratified by time since vaccination in IIC. <sup>b</sup>Calculated per 1000 person-days. <sup>c</sup>Adjusted for age, sex, income, employed, disability, obesity, current smoking, residence, Charleson comorbidity index, COVID-19 within 6 months prior to index date. aHR, adjusted hazard ratio; IC, immunocompromised; IIC, individuals who are immunocompromised; IR, incidence rate.

For those who received a third dose of NVX-CoV2373, the IR (95% CI) for any SARS-CoV-2 infection was 2.19 (2.10–2.29) in both the IIC and non-IC groups within the 30-day risk window (**Table 3**). The IRs (95% CI) for severe infection in the IIC and non-IC groups in this period were 0.01 (0.01–0.02) and 0.01 (0.00–0.01), respectively. Within the 180-day risk window for the third-dose group, the IR (95% CI) for any SARS-CoV-2 infection in the IIC group was 1.08 (1.05–1.11) and 1.04 (1.01–1.07) in the non-IC group; the IRs for severe infection over the 180-day period were 0.01 (0.00–0.01) in both groups.

For those individuals who received a fourth dose of NVX-CoV2373, the IR (95% CI) within the 30-day risk window for any SARS-CoV-2 infection was 0.78 (0.76–0.80) in the IIC group and 0.76 (0.74–0.79) in the non-IC group and were 0.00 (0.00–0.01) in both groups for severe infection (**Table 3**). Within the 180-day risk window, the IRs (95% CI) for any SARS-CoV-2 infection were 0.68 (0.67–0.69) and 0.66 (0.65–0.67) for the IIC and non-IC groups, respectively, and for severe infection were 0.00 (0.00–0.00) in both groups.

### Infection risk comparison among all IIC and non-IC individuals

Within the 30-day risk window after a primary series of NVX-CoV2373, any (aHR=1.04 [95% CI: 0.96–1.12] or severe (0.72 [0.37–1.38]) SARS-CoV-2 infection did not significantly differ between groups of IIC and non-IC individuals (**Table 3**; **Figure 2**). Through the 180-day risk window after a primary series, there was a significant and modestly higher risk (10%) for IIC, compared to non-IC individuals for any SARS-CoV-2 infection (aHR=1.10 [95% CI: 1.06– 1.14]), but not for severe infection (0.76 [0.52–1.12]). In this same window, there was a slightly increased significant risk of any infection post third dose (aHR=1.05 [95% CI: 1.01–1.09]) and fourth dose (1.03 [1.02–1.05]) but no significant differences for severe infection. Overall, there was a slightly higher and significant risk of any SARS-CoV-2 infection in the fourth-dose group, across all risk windows.

**Figure 2:**
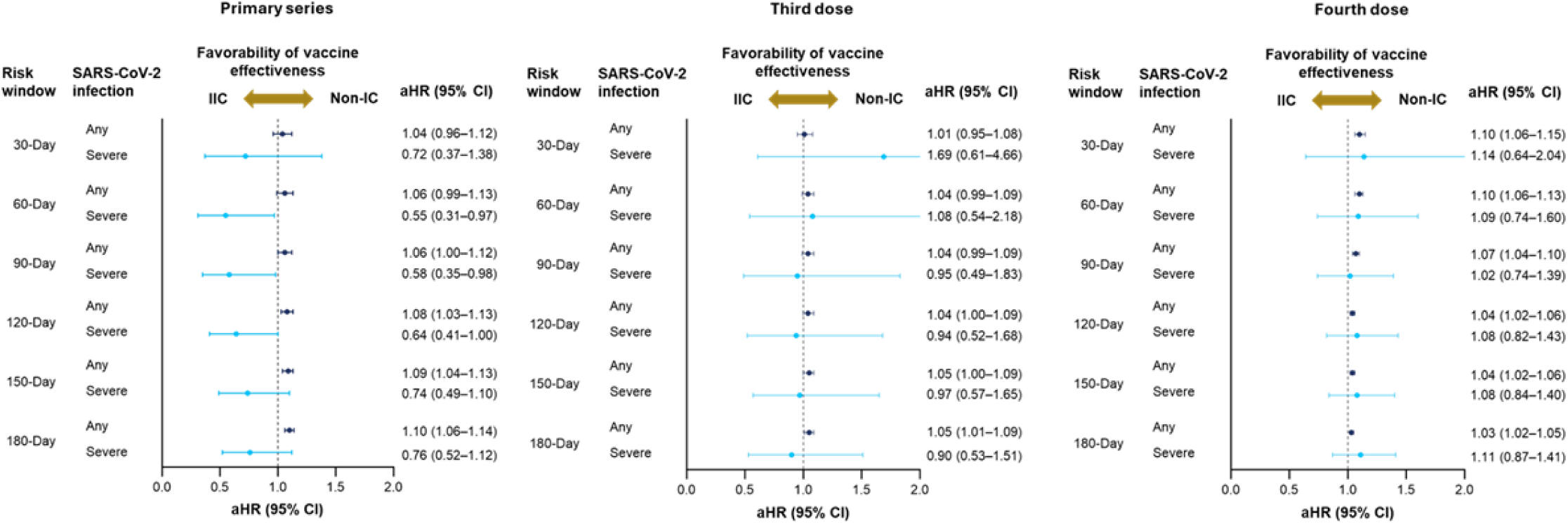
Risk of any or severe SARS-CoV-2 infection after a primary series of NVX-CoV2373 or a third or fourth COVID-19 vaccination among IIC is displayed as adjusted hazard ratios (aHRs) with 95% confidence intervals (CI). Risk windows occurred in 30-day increments and began 14 days after the primary series and 7 days after the third or fourth dose. aHR, adjusted hazard ratio; IIC, individuals who are immunocompromised; Non-IC, individuals who are not immunocompromised.

### Evaluation of residual bias/confounding using the recently vaccinated period

The odds of any SARS-CoV-2 infection in the first 5 days after receipt of NVX-CoV2373 were slightly higher for IIC compared to non-IC individuals in the primary series group, and the difference was statistically significant (aOR=1.21 [95% CI: 1.01–1.45]; **Table S1**). In this early period post vaccination, there was no significant difference in risk of any SARS-CoV-2 infection for IIC compared to non-IC individuals (aOR [95% CI]) in neither the third-dose group (1.13 [0.93–1.39]) nor in the fourth-dose group (1.09 [0.97–1.23]).

### Sensitivity analysis of infection risk in subsets of IIC

Risk estimates for the 180-day risk window were stratified by the type of IIC (i.e., rheumatologic/inflammatory disorder, solid organ malignancy, hematologic malignancy, and organ or stem cell transplant), to assess a subset of individuals considered to have a more severe immunocompromising condition (**Table S2**).

A significantly higher risk of any SARS-CoV-2 infection was observed for individuals with a rheumatologic/inflammatory disorder across the primary series (aHR=1.14 [95% CI: 1.09–1.19]), third-dose, and fourth-dose groups. Among individuals with a solid organ malignancy, there was no significant difference in risk of any SARS-CoV-2 infection, compared with non-IC individuals, after a primary series, third dose, or fourth dose of NVX-CoV2373. For individuals with a hematologic malignancy, risk estimates (aHR [95% CI]) were generally comparable to non-IC individuals; however, there was a significantly higher risk observed for any SARS-CoV-2 infection in the primary series group (1.27 [95% CI: 1.06–1.51]). IIC due to organ or stem cell transplant had a significantly higher risk of any SARS-CoV-2 infection in the primary series (aHR=1.30 [95% CI: 1.06–1.60]) and fourth-dose groups compared with non-IC. Risk estimates for severe SARS-CoV-2 infection were also calculated for each IIC subgroup and are shown in Table S2.

## Discussion

These findings indicate that NVX-CoV2373 provides relatively similar protection against any and severe SARS-CoV-2 infection in both IIC and non-IC individuals across the primary series and third and fourth doses. Overall, risk of any infection among IIC was a maximum of 10% higher, compared to the non-IC group.

Although risk differences were statistically significant in some instances, the clinical relevance of these marginal differences is not clear. Residual bias assessment of infection in the first five days post vaccination indicated significantly higher odds of any SARS-CoV-2 infection in IIC, compared to their non-IC counterparts after a primary series of NVX-CoV2373. This suggests that IIC might have had a higher baseline risk for medically attended SARS-CoV-2 infection, or could have been more likely to seek healthcare, compared to those without an immunocompromising condition. The group of IIC is very heterogeneous; therefore, a sensitivity analysis was performed to gather more information by examining infection in the 180-day risk window, in subsets of IIC with more severe immunocompromising conditions. Severe SARS-CoV-2 infections were assessed in the sensitivity analysis; however, the small sample size and low number of severe infections confound interpretation of these results.

The largest subgroup included in the sensitivity analysis comprised IIC with a rheumatologic/inflammatory disorder (56.0%). A small, but significantly increased risk of any SARS-CoV-2 infection was observed for all dose groups within this subgroup, compared to non-IC individuals; however, this subset of individuals is comprised of a heterogenous group of conditions and immune-modulating medications [18], which require a more in-depth analysis. Risk of SARS-CoV-2 infection was relatively similar between IIC with solid organ malignancy and the non-IC group across each of the dose groups. One reason for these comparable outcomes may be potentially less severe immunosuppression among those with solid organ malignancy, compared with other subgroups of IIC. Specifically, immune modulation is localized to the tumor microenvironment from targeted therapies [19] and chemotherapy treatment is shorter in duration (e.g., weeks/months) compared with life-long treatments for other conditions in IIC [20–22].

There was a statistically significant increase in risk (27%) of any SARS-CoV-2 infection in IIC with a hematologic malignancy compared with non-IC individuals in the primary series group. Considering that hematologic malignancies affect the bone marrow and production of lymphocytes, as does aggressive myelosuppressive treatment [23], these results are not surprising. Notably, in the OCTAVE-DUO study, participants with a hematologic malignancy who had low responses to a COVID-19 vaccine primary series were vaccinated with NVX-CoV2373 and had serological responses similar to those observed in healthy recipients of a primary series [24]. The results from the OCTAVE-DUO study, taken together with the general lack of significantly increased infection risk in the sensitivity analysis for this study, NVX-CoV2373 demonstrated relative effectiveness in those with a hematologic malignancy.

Although only 0.5% of the IIC population in this study were individuals having undergone an organ or stem cell transplant, this subgroup was included in the sensitivity analysis based on their long-term/life-long use of immunosuppressive medications [25,26]. Results from this analysis indicate that individuals who have undergone an organ or stem cell transplant have significantly higher risk for any infection (∼30% in the primary series and fourth-dose groups) compared with non-IC individuals. Despite the increased risk compared to non-IC individuals reported here, additional doses of COVID-19 vaccines are likely to afford some level of protection for this subgroup. For example, a study that included 30 kidney transplant recipients who had inadequate immune responses to four prior doses of mRNA-based COVID-19 vaccines reported seroconversion in 44% and robust immune responses in 17% of the population after receipt of a study dose of NVX-CoV2373 [27].

The population of IIC in this study encompasses conditions observed in the global population; however, the heterogeneity of conditions included can be considered a limitation with varying rVE among the different subgroups. The sensitivity analysis aimed to address concerns over heterogeneity by investigating outcomes in subsets of individuals considered to be severely immunocompromised; however, the low number of severe SARS-CoV-2 infections that occurred might have impacted the precision of analyzing that related risk within the condition subgroups, as well as in the overall population. Information on concomitant medications were unavailable, and these could have an impact on vaccine response and disease severity [28,29]. Propensity score matching and adjustments based on recent vaccination provided a balanced comparison between IIC and non-IC groups; however, there is the potential for additional confounders. Finally, this analysis includes only the prototype vaccine to ancestral SARS-CoV-2 (Wuhan), with data collection including a period when variants were in circulation [16], and data are unavailable for which strains caused documented infections.

## Conclusion

Overall, in this analysis of real-world data from individuals in South Korea, NVX-CoV2373 provided protection against any and severe SARS-CoV-2 infection for IIC, largely to a similar degree when compared with non-IC individuals. Protection was observed with both a homologous primary series and homologous or heterologous third and fourth doses of NVX-CoV2373, as well as across a variety of immunocompromising conditions.

## ABBREVIATIONS

aHR: adjusted hazard ratio;
aSD: absolute standardized difference;
IC: immunocompromised;
ICU: intensive care unit;
IIC: individuals who are immunocompromised;
IR: incidence rate;
rS: recombinant spike;
rVE: relative vaccine effectiveness;
VE: vaccine effectiveness.

## Author contributions

EG, S-AC, MHG, MV, MDR, and YJC conceptualized the trial and contributed to its design.

EG, S-AC, KK, and EB were involved in the extraction and analysis of data.

EG, S-AC, KK, EB, MHG, JF, MV, MDR, and YJC were involved in the interpretation of data.

EG, S-AC, KK, EB, and YJC verified the data.

EG, S-AC, KK, EB, MHG, JF, MV, MDR, and YJC drafted the manuscript.

EG, S-AC, and KK performed the statistical analyses.

S-AC, MHG, JF, MDR, and YJC supervised the study.

All authors critically reviewed and approved the manuscript content.

## Funding

Funding was provided by Novavax, Inc.

## Competing interests

EG, SAC, KK, EB, and YJC are study investigators and do not have any conflicts of interest to report. JF, MHG, and MDR are employees of Novavax, Inc. and may own stock. MV was a salaried employee of Novavax, Inc. and may own stock.

## Ethics approval

Data from the K-COV-N database were anonymized. As this was a retrospective review, no specific ethics approvals were required.

## Acknowledgments

Medical writing and editorial support were provided by Kelly M. Fahrbach, PhD, CMPP, and Ebenezer M. Awuah-Yeboah, BS, of Ashfield MedComms (New York, USA), an Inizio company, supported by Novavax, Inc.

## Data availability

The data used in this study are from the K-CoV-N cohort, which is not publicly available. Researchers can apply for access to these data through the National Health Insurance Service (NHIS) and Korea Disease Control and Prevention (KDCA) data linkage system. Applications can be submitted via the Health Care Data Linkage (HCDL) website (https://hcdl.mohw.go.kr/) and are subject to review by the relevant authorities.

## Notes

### Author Declarations

The study protocol was approved by the Korea University Institutional Review Board (IRB No. 2023AN0124).

